# COVID-19, smoking, vaping and quitting: A representative population survey in England

**DOI:** 10.1101/2020.06.29.20142661

**Authors:** Harry Tattan-Birch, Olga Perski, Sarah Jackson, Lion Shahab, Robert West, Jamie Brown

## Abstract

**Aims:** To explore 1) associations between suspected SARS-CoV-2 infection, hand washing, smoking status, e-cigarette use, and nicotine replacement therapy (NRT) use and 2) whether COVID-19 has prompted smoking and vaping quit attempts, and more smoking inside the home.

**Design:** Cross-sectional household surveys of a representative sample of the population in England from April–May 2020.

**Participants:** The sample included 3,285 adults aged ≥18 years.

**Measurements:** Participants who reported they definitely or think they had coronavirus were classified as having a suspected SARS-CoV-2 infection. Participants were asked how often they wash their hands after returning home, before eating, before preparing foods or before touching their face. They were also asked whether, due to COVID-19, they had i) attempted to quit smoking, ii) attempted to quit vaping, and iii) changed the amount they smoke inside the home.

**Findings:** Odds of suspected SARS-CoV-2 infection were significantly greater among current smokers (20.9%, adjusted odds ratio [OR_adj_]=1.34, 95% confidence interval [CI]=1.04–1.73) and long-term (>1-year) ex-smokers (16.1%, OR_adj_=1.33, 95%CI=1.05–1.68) than never smokers (14.5%). Recent (<1-year) ex-smokers had non-significantly greater odds of suspected infection (22.2%, OR_adj_=1.50, 95%CI=0.85–2.53, Bayes factor= 0.55–1.17). Bayes factors indicated there was sufficient evidence to rule out large differences in suspected SARS-CoV-2 infection by NRT use and medium differences by e-cigarette use. With the exception of hand washing before face touching, engagement in hand washing behaviours was high (>85%) regardless of nicotine use. A minority (12.2%) of past-year smokers who made a quit attempt in the past three months were triggered by COVID-19, and approximately one-in-ten current e-cigarette users reported attempting to quit vaping because of COVID-19. Most people reported smoking the same amount inside the home.

**Conclusions:** In a representative sample of the adult population in England, current smokers and long-term ex-smokers had higher odds of suspected SARS-CoV-2 infection than never smokers, but there were no large differences by NRT or e-cigarette use. In general, engagement in hand washing was high regardless of nicotine or tobacco use. A minority of past-year smokers and current e-cigarette users, respectively, attempted to quit smoking/vaping due to COVID-19.

## Introduction

COVID-19 is a respiratory disease caused by the SARS-CoV-2 virus.^1^ Tobacco smoking was considered an *a priori* risk factor for SARS-CoV-2 infection and poor COVID-19 disease outcomes: current and former smoking is known to increase the risk of respiratory viral and bacterial infection and adverse disease outcomes compared with never smoking.^2,3^ Behavioural factors involved in both smoking and vaping, such as regular hand-to-mouth movements, may increase viral infection and transmission if performed without accompanying protective behaviours like hand washing.^4^ However, early descriptive epidemiology from the ongoing pandemic produced surprising results. Limited, mixed-quality evidence suggests lower than expected smoking rates among those testing positive for SARS-CoV-2 infection and those hospitalised with COVID-19.^5,6^ This has led to the hypothesis that nicotine may protect against a hyper-inflammatory response to SARS-CoV-2 infection,^7,8^ thus preventing adverse outcomes such as hospitalisation with COVID-19 disease. Alternatively, the lower than expected smoking rates may reflect smokers being less likely to become infected due to an unexpected interaction between nicotine and ACE2 receptors, or may just be an artefact of various measurement and sampling issues.^5^ Our aim is to use a large, representative population survey with reliable assessment of smoking status to estimate rates of self-reported SARS-CoV-2 infection in relation to smoking status.

The early literature has focussed on smoking, SARS-CoV-2 infection and COVID-19 disease. However, comparisons of rates of symptomatic SARS-CoV-2 infection between smokers, ex-smokers and non-smokers are a poor test of the nicotine hypothesis. Other components in tobacco smoke could lead to greater symptom severity, which could obscure the hypothesised protective effect of nicotine. Electronic cigarettes (e-cigarettes) and nicotine replacement therapy (NRT) have lower toxicity than cigarettes.^9,10^ Therefore, an assessment of the use of these products provides a better test of a potential protective effect of nicotine. Despite this, few studies so far in the pandemic have measured the use of e-cigarettes or NRT.^11^ In addition, most studies have not been able to rely on representative sampling and have failed to distinguish between current smokers, recent ex-smokers, long-term ex-smokers and never smokers.^5^ Our secondary aim is therefore to estimate rates of SARS-CoV-2 infection by use of alternative nicotine products in a representative population survey.

Hand washing is an important behaviour in reducing the transmission of infectious diseases through the population, especially for those that are primarily spread through respiratory droplets.^12^ Because of the repeated hand-to-mouth movements involved in smoking and vaping, current smokers and e-cigarette users may be less likely to regularly wash their hands before touching their face. Additionally, personality factors, such as lower risk aversion,^13^ could lead to reduced hand washing among tobacco and nicotine users. These behavioural factors could explain differences in rates of SARS-CoV-2 infection by smoking status, e-cigarette use or NRT use. Therefore, in this study, we measured associations between self-reported engagement in hand washing behaviours and use of these products.

In the absence of clear evidence as to whether smoking is a risk factor for SARS-CoV-2 infection or hospitalisation, public health messaging has focused on general benefits of quitting tobacco smoking.^14,15^ Some organisations, such as the American Lung Association,^16^ have also urged people to stop using e-cigarettes (“vaping”), claiming that doing so may reduce their risk of developing severe COVID-19 symptoms — despite little evidence to support this. The SARS-CoV-2 pandemic may therefore have prompted smokers and e-cigarette users to attempt to stop in greater numbers. Social distancing measures, introduced in England in March 2020, have limited people to leave their home only for food, healthcare, exercise and essential work.^17^ These measures may have increased the amount people smoke inside their home, which would be especially damaging because school closures mean children are housebound and at risk of exposure to second-hand smoke.

The COVID-19 pandemic has exacerbated heath inequalities in England, with the highest mortality rates found among people working in manual, routine and service occupations^18^ and in black and ethnic minority groups.^19^ In addition, those living in deprived areas are the most likely to be diagnosed with COVID-19 and have worse outcomes when hospitalised.^20^ Because smoking and vaping prevalence is highest among people from disadvantaged groups,^21^ the impact these behaviours have on SARS-CoV-2 infection may be more pronounced in these groups. In this study, we will use a representative population sample of adults in England to estimate:

1. associations between suspected SARS-CoV-2 infection and smoking status, e-cigarette use and NRT use;
2. associations between hand washing and smoking status, e-cigarette use and NRT use;
3. the proportion of smokers and e-cigarette users who have attempted to quit product use because they are motivated by the pandemic;
4. the proportion of smokers, with or without children in their household, who are smoking more, less, or the same amount inside their home; and
5. whether the above associations vary by socioeconomic status (SES).

## Methods

### Design

This study is part of the ongoing Smoking Toolkit Study (STS), a monthly cross-sectional survey in England. It provides detailed information on smoking, vaping and NRT use. It recruits around 1700 participants per month using a combination of random location and quota sampling.^22^ Interviews are performed with one household member until quotas based on factors influencing the probability of being at home (e.g. gender, age, working status) are fulfilled. Comparisons with sales data and other national surveys show that the STS recruits a representative sample of the population in England with regard to key demographic variables, smoking prevalence and cigarette consumption.^22,23^ Data are usually collected through face-to-face computer assisted interviews. However, social distancing restrictions under the COVID-19 pandemic mean that data from April and May 2020 were collected via telephone but relied on the same combination of random location and quota sampling, and weighting approach. Ethical approval was provided by the UCL Research Ethics Committee (0498/001).

### Study sample

Adults (≥18 years) who were interviewed in April and May 2020, the first survey waves to include questions about COVID-19.

### Measures

#### Smoking status

Participants who reported currently smoking tobacco of any kind were considered *current smokers*. Those who reported having stopped smoking within the last year were considered *recent ex-smokers*. Those who reported having stopped smoking more than a year ago were considered *long-term ex-smokers*. All others were considered *never smokers*. Never smokers were used as the reference category when calculating odds ratios.

#### E-cigarette and NRT use

Participants were asked which products they are currently using to cut down the amount they smoke, in situations when they are not allowed to smoke, to help them stop smoking, or for any other reason at all. Those who reported currently using e-cigarettes were considered *current e-cigarette users* and those who reported currently using NRT (e.g. gum, patches, lozenges) were considered *current NRT users*.

#### Suspected SARS-CoV-2 infection

Participants were told that the symptoms of coronavirus are high temperature/fever or a new, continuous cough. They were then asked which of the following statements best applies to them: (1) I definitely have coronavirus, (2) I think I have coronavirus, (3) I definitely had coronavirus, (4) I think I had coronavirus, (5) I do not have or think I have had coronavirus, (6) Don’t know, and (7) Prefer not to say. Those who responded that they “definitely” or “think they” have/had coronavirus (1 – 4) were labelled as having a *suspected SARS-CoV-2 infection*.

#### Hand washing

Participants were asked how frequently, in the last week, they (i) washed their hands when they got home after being out, (ii) washed their hands before they ate, (iii) washed their hands before they prepared food, and (iv) touched their eyes, nose or mouth without washing their hands first. For all questions, participants could respond always, most of the time, about half the time, less than half the time, never or don’t know. Responses were grouped into two categories: engaging in the behaviour the majority (most/always) of the time, coded 1, and all others, coded 0. For question (i), participants could respond that they have not left the house, and for question (ii), that they have not prepared food in the last week. Individuals who gave these responses were considered protected against infection, with responses coded as 1. Responses from question (iv) were reverse coded to reflect the protective behaviour of washing hands before touching eyes, nose or mouth.

#### Smoking quit attempt

Past-year smokers who reported that they had made a quit attempt in the last 3 months were asked if their attempt was triggered by (i) the COVID-19 outbreak or (ii) concern about future health problems.

#### Vaping quit attempt

Current e-cigarette users were asked if they were trying to quit vaping completely because of the COVID-19 outbreak.

#### Smoking inside the home

Smokers were asked whether, in the past month, they had been smoking more, the same amount or fewer cigarettes inside their home.

#### Children in household

Smokers were asked the number of children they had in their household. Responses were dichotomised as ≥1 and 0.

#### SES

Occupation-based social grade^24^ was used as our measure of SES, with responses dichotomised into ABC1 (managerial, professional and clerical workers) and C2DE (manual and casual workers, state pensioners and unemployed).

#### Potential confounders

In addition to SES, we included information on sex, age and region (London, North, Central, South).

### Analysis

All analyses were conducted in R version 3.5.^25^ The pre-registered analysis plan is available on the Open Science Framework (https://osf.io/vb9fm/). We made one amendment, which was to include data from May 2020 when this became available. Results following the original plan based on April 2020 data are available online (https://osf.io/7ehx6/). Alpha was set to 5%. Where there were nonsignificant results when assessing analysis 1 (A1), Bayes factors (BFs) were used to distinguish inconclusive data from evidence for no large, medium or small effect.^26,27^ Large effects in either direction were defined as having an odds ratio (OR) = 4 or for lower estimates ¼, based on evidence that current smoking rates among patients hospitalised for COVID-19 in China were four times lower than would be expected from the smoking prevalence in the country.^6^ We defined a medium effect size as OR = 2 or ½ and a small effect size as OR = 1.5 or 2/3.

#### A1

Logistic regression was used to estimate the association between suspected SARS-CoV-2 infection and (i) smoking status, (ii) e-cigarette use and (iii) NRT use, with and without adjustment for potential confounders. We repeated the above analysis but included the interaction between SES and (i) through (iii) as an explanatory variable.

#### A2

Similarly, for each hand washing behaviour, we used logistic regression to estimate the association between engaging in the behaviour and (i) smoking status, (ii) e-cigarette use and (iii) NRT use, with and without adjustment for potential confounders.

#### A3

We report the proportion of (i) past-year smokers and (ii) e-cigarette users who attempted to quit because of the outbreak, before and after stratifying by SES. For smokers, we also compared this to the proportion who attempted to quit because they were concerned about their future health.

#### A4

We also report the proportion of smokers, with or without children in their household, who were smoking more, less or the same amount inside their home. Logistic regression was used to estimate the association of smoking more (vs. less/about the same amount) inside the home with children in household, SES, and the interaction between SES and children in the household.

## Results

Of the 3,285 participants interviewed, 3,179 (96.8%) provided complete data on all questions required for the current analyses. Sample characteristics are shown in Table 1. Of the analytic sample, 1,804 (56.7%) were never smokers, 834 (26.2%) were long-term ex-smokers, 72 (2.3%) were recent ex-smokers, and 469 (14.8%) were current smokers. In addition, 192 (6.0%) were current e-cigarette users and 88 (2.8%) were current NRT users.

**Table 1.** Sample characteristics.

|  | Analytical<br>sample (n =<br>3,179) |
| --- | --- |
| Sex |  |
| Women | 1718 (54.0%) |
| Age |  |
| Mean (sd) | 52.4 (17.6) |
| Region |  |
| London | 462 (14.5%) |
| North | 938 (29.5%) |
| Central | 951 (29.9%) |
| South | 828 (26.0%) |
| SES |  |
| ABC1 | 1969 (61.9%) |
| C2DE | 1210 (38.1%) |

### A1: Suspected SARS-CoV-2 infection

Of the sample, 16.0% (95% confidence interval [CI] = 14.8%–17.4%) reported a suspected SARS-CoV-2 infection. Table 2 shows the prevalence of suspected SARS-CoV-2 infection by smoking status, e-cigarette and NRT use. In adjusted analyses, current smokers and long-term ex-smokers had significantly greater odds of suspected SARS-CoV-2 infection compared with never smokers. There were no other significant differences by smoking status, e-cigarette use or NRT use, or significant interactions between product use and SES (Supplementary Table 1). BFs indicated that there was insufficient evidence to rule out small, medium or large differences in suspected SARS-CoV-2 infection between recent ex-smokers and never smokers (Supplementary Table 2). There was sufficient evidence to rule out (1) large unadjusted and adjusted associations with NRT use and (2) medium and large adjusted associations with e-cigarette use.

**Table 2:** Associations of suspected SARS-CoV-2 infection with smoking status, e-cigarette use and NRT use.

| Variable | N (% [95% CI]) | OR (95% CI) | p | OR<sub>adj</sub> (95% CI) | p |
| --- | --- | --- | --- | --- | --- |
| Smoking status |  |  |  |  |  |
| Never | 261 (14.5% [12.9%-16.2%]) | - | - | - | - |
| Long-term ex | 135 (16.1% [13.8%-18.8%]) | 1.18 (0.94-1.48) | .16 | 1.33 (1.05-1.68) | .02 |
| Recent ex | 16 (22.2% [14.2%-33.1%]) | 1.61 (0.92-2.69) | .08 | 1.50 (0.85-2.53) | .14 |
| Current smoker | 98 (20.9% [17.5%-24.8%]) | 1.44 (1.13-1.84) | <.01 | 1.34 (1.04-1.73) | .02 |
| E-Cigarette use |  |  |  |  |  |
| No current use | 468 (15.7% [14.4%-17.0%]) | - | - | - | - |
| Current use | 42 (21.9% [16.6%-28.2%]) | 1.30 (0.91-1.81) | .14 | 1.00 (0.68-1.43) | .99 |
| NRT use |  |  |  |  |  |
| No current use | 494 (16.0% [14.7%-17.3%]) | - | - | - | - |
| Current use | 16 (18.2% [11.5%-27.5%]) | 1.00 (0.55-1.68) | .99 | 0.82 (0.44-1.41) | .49 |

### A2: Hand washing

Of participants, 95.6% (95.0%-96.4%) reported washing their hands most of the time after coming home, 97.1% (96.5–97.6%) reported washing their hands most of the time before they prepared food, 87.2% (86.0%–88.3%) reported washing their hands most of the time before they ate, and 71.3% (69.7%–72.9%) reported not touching their face without washing their hands at least half of the time.

Compared with never smokers, recent ex-smokers had significantly lower odds of reporting washing their hands before touching their face the majority of the time, both before and after adjustment for potential confounding variables (see Table 3). All other associations of hand washing with smoking status, e-cigarette use and NRT use were non-significant.

**Table 3:** Associations between hand washing and smoking status, e-cigarette use and NRT use.

| Hand washing context | N (% [95% CI]) | OR<sub>un</sub> (95% CI) | p | OR<sub>adj</sub> (95% CI) | p |
| --- | --- | --- | --- | --- | --- |
| **After coming home** |  |  |  |  |  |
| Smoking status |  |  |  |  |  |
| Never | 1728 (95.8% [94.8%-96.6%]) | - | - | - | - |
| Long-term ex | 803 (96.3% [94.8%-97.4%]) | 1.25 ( 0.82- 1.94) | .31 | 1.20 ( 0.78- 1.89) | .41 |
| Recent ex | 67 (93.1% [84.8%-97.0%]) | 0.67 ( 0.31- 1.80) | .37 | 0.76 ( 0.34- 2.06) | .55 |
| Current smoker | 446 (95.1% [92.7%-96.7%]) | 0.94 ( 0.61- 1.48) | .78 | 1.10 ( 0.71- 1.76) | .68 |
| E-cigarette use |  |  |  |  |  |
| No current use | 2859 (95.7% [94.9%-96.4%]) | - | - | - | - |
| Current use | 185 (96.4% [92.7%-98.2%]) | 1.61 ( 0.79- 3.93) | .24 | 1.87 ( 0.88- 4.68) | .14 |
| NRT use |  |  |  |  |  |
| No current use | 2959 (95.7% [95.0%-96.4%]) | - | - | - | - |
| Current use | 85 (96.6% [90.5%-98.8%]) | 1.50 ( 0.55- 6.15) | .50 | 1.73 ( 0.62- 7.25) | .37 |
| **Before preparing food** |  |  |  |  |  |
| Smoking status |  |  |  |  |  |
| Never | 1758 (97.5% [96.6%-98.1%]) | - | - | - | - |
| Long-term ex | 808 (96.9% [95.5%-97.9%]) | 0.66 ( 0.41- 1.10) | .10 | 0.71 ( 0.43- 1.18) | .18 |
| Recent ex | 69 (95.8% [88.5%-98.6%]) | 0.60 ( 0.22- 2.40) | .39 | 0.55 ( 0.20- 2.23) | .32 |
| Current smoker | 452 (96.4% [94.3%-97.7%]) | 0.67 ( 0.39- 1.18) | .15 | 0.65 ( 0.37- 1.17) | .14 |
| E-cigarette use |  |  |  |  |  |
| No current use | 2899 (97.1% [96.4%-97.6%]) | - | - | - | - |
| Current use | 188 (97.9% [94.8%-99.2%]) | 1.64 ( 0.67- 5.48) | .34 | 2.00 ( 0.79- 6.83) | .20 |
| NRT use |  |  |  |  |  |
| No current use | 3002 (97.1% [96.5%-97.7%]) | - | - | - | - |
| Current use | 85 (96.6% [90.5%-98.8%]) | 1.11 ( 0.37- 5.59) | .87 | 1.40 ( 0.45- 7.21) | .62 |
| **Before eating** |  |  |  |  |  |
| Smoking status |  |  |  |  |  |
| Never | 1583 (87.7% [86.2%-89.2%]) | - | - | - | - |
| Long-term ex | 720 (86.3% [83.8%-88.5%]) | 0.87 (0.68-1.12) | .27 | 0.88 (0.68-1.14) | .33 |
| Recent ex | 62 (86.1% [76.3%-92.3%]) | 1.03 (0.55-2.13) | .94 | 0.94 (0.50-1.98) | .87 |
| Current smoker | 407 (86.8% [83.4%-89.5%]) | 0.98 (0.74-1.32) | .91 | 0.94 (0.70-1.28) | .71 |
| E-cigarette use |  |  |  |  |  |
| No current use | 2604 (87.2% [85.9%-88.3%]) | - | - | - | - |
| Current use | 168 (87.5% [82.1%-91.5%]) | 1.20 (0.79-1.90) | .42 | 1.22 (0.78-1.98) | .40 |
| NRT use |  |  |  |  |  |
| No current use | 2698 (87.3% [86.1%-88.4%]) | - | - | - | - |
| Current use | 74 (84.1% [75.0%-90.3%]) | 1.06 (0.59-2.09) | .86 | 1.10 (0.59-2.21) | .78 |
| **Before touching face** |  |  |  |  |  |
| Smoking status |  |  |  |  |  |
| Never | 1316 (72.9% [70.9%-74.9%]) | - | - | - | - |
| Long-term ex | 576 (69.1% [65.8%-72.1%]) | 0.88 (0.73-1.06) | .19 | 0.86 (0.71-1.04) | .11 |
| Recent ex | 42 (58.3% [46.8%-69.0%]) | 0.55 (0.36-0.87) | <.01 | 0.57 (0.37-0.91) | .02 |
| Current smoker | 334 (71.2% [67.0%-75.1%]) | 0.89 (0.72-1.10) | .28 | 0.94 (0.76-1.16) | .54 |
| E-cigarette use |  |  |  |  |  |
| No current use | 2128 (71.2% [69.6%-72.8%]) | - | - | - | - |
| Current use | 140 (72.9% [66.2%-78.7%]) | 1.12 (0.83-1.53) | .47 | 1.30 (0.94-1.81) | .12 |
| NRT use |  |  |  |  |  |
| No current use | 2204 (71.3% [69.7%-72.9%]) | - | - | - | - |
| Current use | 64 (72.7% [62.6%-80.9%]) | 1.02 (0.66-1.63) | .92 | 1.12 (0.71-1.82) | .63 |

Adjusted analyses included terms for SES, sex, age and region. In addition, smoking status was added as a covariate when estimating associations between the outcome and e-cigarette use and NRT use.

### A3: Quitting

There were 49 past-year smokers who attempted to quit in the past three months, 6 (12.2%, 5.7%-24.2%) of whom reported attempting to quit smoking due to COVID-19. The proportion who reported attempting to quit smoking due to COVID-19 was similar across advantaged (n=3, 13.0%, 4.5%-32.1%) and disadvantaged (n=3, 11.5%, 4.0%-29.0%) groups. In comparison, 20 (40.8%, 28.2%-54.8%) reported being motivated to quit smoking because of future health concerns, again with similar proportions from advantaged (n=8, 34.8%, 18.8%-55.1%) and disadvantaged (n=12, 46.2%, 28.7%-64.5%) groups. Of the 170 current e-cigarette users surveyed, 19 (11.2%, 7.3%-16.8%) attempted to quit vaping because of COVID-19. This was similar across SES: 12 (12.6%, 7.4%-20.8%) of the 95 advantaged current e-cigarette users attempted to quit vaping because of COVID-19, compared with 7 (9.3%, 4.6%-18.0%) of the 75 disadvantaged users.

### A4: Smoking inside the home

Of the 469 current smokers, 223 (47.5%, 43.1%-52.1%) reported smoking the same number of cigarettes inside their home in the past month, 109 (23.2%, 19.6%-27.3%) reported smoking fewer cigarettes inside their home, 112 (23.9%, 20.2%-27.9%) reported smoking more cigarettes inside their home and 15 (3.2%, 1.9%-5.2%) didn’t know. As is shown in Table 4, odds of smoking more inside the home did not significantly differ between advantaged and disadvantaged smokers or between those with or without children in the home. In addition, there was no significant interaction between SES and children in the home (OR = 1.79, 0.73-4.33, *p =* .20).

**Table 4:** Associations between smoking more inside the home, SES and children in the home.

| Variable | N (%[95% CI]) | OR (95% CI) | p |
| --- | --- | --- | --- |
| SES |  |  |  |
| ABC1 | 46 (20.4% [15.7%-26.2%]) | - | - |
| C2DE | 66 (27.0% [21.9%-32.9%]) | 1.42 (0.94 - 2.16) | .10 |
| Children in home |  |  |  |
| Children | 30 (20.5% [14.8%-27.8%]) | - | - |
| No children | 82 (25.4% [20.9%-30.4%]) | 1.29 (0.85 - 1.98) | .23 |

## Discussion

In this representative sample of adults in England, current smokers and long-term ex-smokers had significantly greater odds of suspected SARS-CoV-2 infection compared with never smokers after controlling for a number of potential confounding variables. There was sufficient evidence to rule out a large association of suspected SARS-CoV-2 infection with NRT use, and a medium association with e-cigarette use. Compared with never smokers, recent ex-smokers had significantly lower odds of washing their hands before touching their face. Participants reported high engagement in other hand washing behaviours, regardless of smoking status, e-cigarette use or NRT use. 12.2% of past-year smokers who made a recent quit attempt were motivated to do so by COVID-19 and a similar proportion (11.2%) of current e-cigarette users attempted to quit vaping due to COVID-19. Most current smokers reported smoking the same number of cigarettes inside their home, while equal proportions reported smoking more or fewer. No significant differences by SES were observed across analyses.

Prior studies unexpectedly found that smoking rates among those hospitalised for COVID-19 were lower than would be expected from population estimates.^5^ There are at least three possible explanations for the observation: smokers are less likely to become infected, smokers who are infected are less likely to develop disease severe enough to require hospitalisation, or it is an artefact of various measurement or sampling issues. In this study, we found that, compared with never smokers, current smokers and long-term ex-smokers had *greater* odds of suspected SARS-CoV-2 *infection*. Recent ex-smokers also had greater odds of suspected infection, but not significantly so. These findings suggest that one of the two explanations other than infection are likely to account for the lower than expected smoking rates among those hospitalised for COVID-19. However, see below for an alternative interpretation.

In terms of infection, the most parsimonious interpretation of the current findings is that current smokers and long-term ex-smokers are at increased risk of SARS-CoV-2 infection, independent of a variety of sociodemographic factors. This increased risk could be the result of behaviour or biology. A relevant behaviour is that smoking requires repeated hand-to-mouth actions although we found no equivalent association with e-cigarette use, which involves similar behaviours. In terms of biology, smoking has been associated with upregulation of ACE2, the receptor for the virus in the lung, which would provide a mechanism for increased risk. However, other studies argue the relationship between smoking and ACE2 is more complicated and may vary between epithelial or alveolar cells.^28,29^ The alternative explanation is that smokers are more likely to report suspected infection but that this does not reflect an increased infection risk. Instead, these differences could result from smokers and long-term ex-smokers having greater susceptibility to other infections with similar symptoms to COVID-19.3 Consistent with this explanation is that current compared with never smoking is associated with reduced risk of testing positive among people tested in the community.^5,30^ In order to resolve these competing explanations for our observation that smokers had greater odds of reporting suspected infection, future studies need to assess the seroprevalence of SARS-CoV-2 infection by smoking status in large representative randomly selected samples of the population.

Beyond smoking, the current findings are also difficult to reconcile with nicotine being substantially protective of SARS-CoV-2 infection. Bayes factors indicated there was sufficient evidence to rule out large differences in suspected SARS-CoV-2 infection by NRT use and medium differences by e-cigarette use. However, the current study was not able to address the possibility that nicotine may protect against the development of more severe COVID-19 disease among people who become infected.

Protective behaviours, such as hand washing, are important in slowing the transmission of infections like SARS-CoV-2. Here we found that, regardless of smoking status or nicotine use, the majority of respondents (>85%) reported that they wash their hands most of the time after coming home, before preparing food and before eating. Fewer people reported washing their hands most of the time before touching their face, and recent ex-smokers had significantly lower odds than never smokers of reporting doing so. The primary route through which people become infected with SARS-CoV-2 is through cells in the lungs and mucus membranes of the eyes, nose and mouth.^31^ Avoiding face touching without first washing one’s hands could therefore be an important and understudied behaviour to reduce transmission.^12^

Public health bodies have advised smokers that quitting may reduce their risk of severe COVID-19 outcomes.^15^ This advice is based on the observation that despite being hospitalised at lower than expected rates, current smoking is associated with greater disease severity among those who are hospitalised.^5^ This advice has been shared through the news and in social media campaigns, such as “#QuitForCovid” in the UK.^14,32^ Here, we found that, while COVID-19 triggered quit attempts among some smokers in England, far more attempts were triggered by other reasons like future health concerns. It is possible that conflicting news stories, reporting the possibility that smoking is protective against COVID-19,^33,34^ may have partially undermined attempts to encourage people to quit smoking. Some organisations have also advised people to stop vaping, arguing that e-cigarette use makes one susceptible to SARS-CoV-2 infection — despite little evidence to support this.^5,16^ In our sample, one in ten current e-cigarette users attempted to quit vaping due to concerns about COVID-19. These attempts to quit vaping might stem from the deteriorating perceptions about the harms associated with e-cigarette use among smokers in England.^35,36^

There was a concern that social distancing measures might increase the amount people smoke inside their home, especially for disadvantaged individuals who may live in housing without easy access to outdoor smoking areas. Reassuringly, we found that the majority of participants reported smoking the same number of cigarettes inside the home, and equal numbers reported smoking more or fewer cigarettes. There was no significant association between smoking more inside the home and SES or children in the household. However, given our small sample and the wide confidence intervals, we may have lacked sufficient data to detect differences.

This study benefits from using a representative sample of the population in England, including questions about current e-cigarette and NRT use, and distinguishing between never, current, recent ex-smokers and long-term ex-smokers. However, there were a number of limitations. First, for some comparisons, sample sizes were small which meant there was substantial uncertainly in estimates. Second, suspected SARS-CoV-2 infection was self-reported and not confirmed with a viral or antibody test. Given that many other infections share symptoms with COVID-19, some participants may have misdiagnosed themselves. In addition, it is likely that most participants with asymptomatic cases did not report being infected. Third, individuals with the most severe cases of COVID-19 were likely censored from our analysis, because they may have died or been hospitalised. Thus, if nicotine reduces risk of severe symptoms, fewer infected nicotine users would be censored from the analysis than non-users, leading to an upward bias in the OR between these groups. However, at its peak, only 0.02% of the UK population were in hospital with COVID-19.^37^ This means censoring likely had a negligible effect on the estimated proportion of participants with suspected SARS-CoV-2 infection, which ranged from 14.5% of never smokers to 22.2% of recent ex-smokers. Fourth, social desirability bias may have led participants to overreport their engagement in hand washing in order to be viewed favourably by their interviewer. Fifth, because reported engagement in hand washing was so high, there may have been a ceiling effect whereby we were unable to detect differences by smoking status, e-cigarette use or NRT use. Finally, only current e-cigarette users were asked whether they had attempted to quit vaping because of concerns about COVID-19, which meant we did not capture those who had already quit vaping.

In conclusion, these results do not support the hypothesis that smoking or nicotine is protective against SARS-CoV-2 infection, with current smokers and long-term ex-smokers reporting significantly greater odds of suspected infection than never smokers and no significant difference in odds of suspected infection by use of e-cigarettes or NRT. With the exception of washing hands before touching one’s face, there was high engagement in hand washing irrespective of nicotine product use. Approximately one in ten current e-cigarette users reported attempting to quit vaping due to COVID-19. As countries begin to relax social distancing measures, it is important to continue tracking how nicotine product use interacts with SARS-CoV-2 infection, and how this pandemic will influence smoking, vaping and quitting behaviours in the population.

## Data Availability

For access to the data please contact the lead author or Prof Jamie Brown.

https://osf.io/vb9fm/

## Competing interests

JB has received unrestricted research funding from Pfizer, who manufacture smoking cessation medications. LS has received honoraria for talks, an unrestricted research grant and travel expenses to attend meetings and workshops from Pfizer, and has acted as paid reviewer for grant awarding bodies and as a paid consultant for health care companies. RW has undertaken consultancy for companies that manufacture smoking cessation medications (Pfizer, J&J, and GSK). All authors declare no financial links with tobacco companies or e-cigarette manufacturers or their representatives.

## Funding

Cancer Research UK fund the Smoking Toolkit Study and the salaries of OP and SJ (C1417/A22962). HTB holds a funded studentship from Public Health England (558585/180737). Funders had no role in the design and conduct of the study; collection, management, analysis and interpretation of the data; preparation, review or approval of the manuscript; and decision to submit the manuscript for publication.

## Supplementary Material

**Supplementary Table 1:**
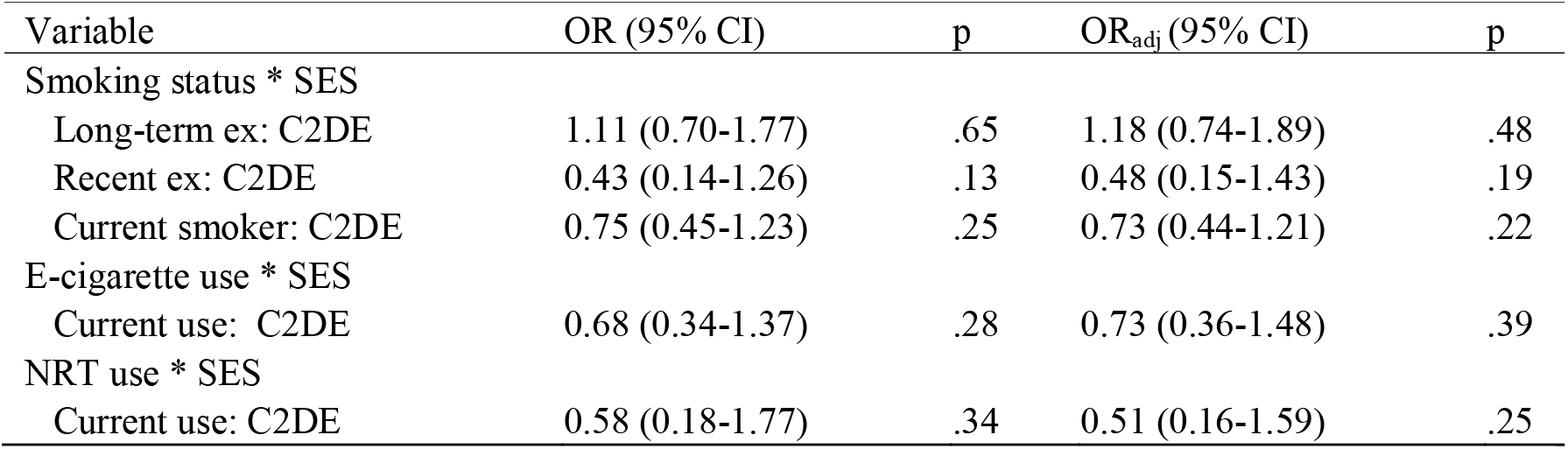
Interaction between SES and smoking status, e-cigarette use and NRT use, when estimating suspected SARS-CoV-2 infection.

| Variable | OR (95% CI) | p | OR<sub>adj</sub> (95% CI) | p |
| --- | --- | --- | --- | --- |
| Smoking status * SES |  |  |  |  |
| Long-term ex: C2DE | 1.11 (0.70-1.77) | .65 | 1.18 (0.74-1.89) | .48 |
| Recent ex: C2DE | 0.43 (0.14-1.26) | .13 | 0.48 (0.15-1.43) | .19 |
| Current smoker: C2DE | 0.75 (0.45-1.23) | .25 | 0.73 (0.44-1.21) | .22 |
| E-cigarette use * SES |  |  |  |  |
| Current use: C2DE | 0.68 (0.34-1.37) | .28 | 0.73 (0.36-1.48) | .39 |
| NRT use * SES |  |  |  |  |
| Current use: C2DE | 0.58 (0.18-1.77) | .34 | 0.51 (0.16-1.59) | .25 |

**Supplementary Table 2:** Bayes factors for non-significant associations between suspected SARS-CoV-2 infection and smoking status, e-cigarette use, and NRT use.

|  | BF<sub>un</sub><br>large | BF<sub>un</sub><br>medium | BF<sub>un</sub><br>small | BF<sub>adj</sub><br>large | BF<sub>adj</sub><br>medium | BF<sub>adj</sub><br>small |
| --- | --- | --- | --- | --- | --- | --- |
| Smoking status |  |  |  |  |  |  |
| Never | - | - | - | - | - | - |
| Long-term ex | - | - | - | - | - | - |
| Recent ex | 0.84 | 1.38 | 1.60 | 0.55 | 0.93 | 1.17 |
| Current smoker | - | - | - | - | - | - |
| E-cigarette use |  |  |  |  |  |  |
| No current use | - | - | - | - | - | - |
| Current use | 0.37 | 0.68 | 0.99 | 0.13 | 0.26 | 0.46 |
| NRT use |  |  |  |  |  |  |
| No current use | - | - | - | - | - | - |
| Current use | 0.20 | 0.38 | 0.57 | 0.26 | 0.48 | 0.68 |

The alternative hypothesis was modelled as a normal distribution centred on zero, with a standard deviation equal to the expected effect size. Large expected effect sizes were set as OR = 4 or 1/4, medium expected effect sizes as OR = 2 or 1/2, small expected effect sizes as OR = 1.5 or 3/2.

**Supplementary Table 3:** Sensitivity analysis estimating the association of suspected SARS-CoV-2 infection with e-cigarette use and NRT use among individuals who were not current smokers.

| Variable | OR (95% CI) | p | OR<sub>adj</sub> (95% CI) | p |
| --- | --- | --- | --- | --- |
| E-Cigarette use |  |  |  |  |
| No current use | - | - | - | - |
| Current use | 1.28 (0.76-2.06) | .32 | 0.96 (0.56-1.59) | .88 |
| NRT use |  |  |  |  |
| No current use | - | - | - | - |
| Current use | 0.89 (0.27-2.25) | .82 | 0.77 (0.23-2.00) | .62 |

Adjusted analyses included terms for SES, sex, age and region. In addition, smoking status was added as a covariate when estimating associations between the outcome and e-cigarette use and NRT use

## Notes

### Clinical Protocols

https://osf.io/s6t9j/

### Author Declarations

Ethical approval was provided by the UCL Research Ethics Committee (0498/001).

